# Incremental diagnostic value of AI-derived coronary artery calcium in ^18^F-flurpiridaz PET Myocardial Perfusion Imaging

**DOI:** 10.1101/2025.07.07.25330013

**Authors:** Orit Barrett, Aakash Shanbhag, Ryan Zaid, Robert J H Miller, Mark Lemley, Valerie Builoff, Joanna X. Liang, Paul B. Kavanagh, Christopher Buckley, Damini Dey, Daniel S. Berman, Piotr J Slomka

**Author notes:** **Address for correspondence:** Piotr J. Slomka, PhD, FACC, Cedars-Sinai Medical Center, 8700 Beverly Blvd, Los Angeles, CA 90048.

## Abstract

**Background:** Positron Emission Tomography (PET) myocardial perfusion imaging (MPI) is a powerful tool for predicting coronary artery disease (CAD). Coronary artery calcium (CAC) provides incremental risk stratification to PET-MPI and enhances diagnostic accuracy. We assessed additive value of CAC score, derived from PET/CT attenuation maps to stress TPD results using the novel 18F-flurpiridaz tracer in detecting significant CAD.

**Methods and Results:** Patients from ^18^F-flurpiridaz phase III clinical trial who underwent PET/CT MPI with ^18^F-flurpiridaz tracer, had available CT attenuation correction (CTAC) scans for CAC scoring, and underwent invasive coronary angiography (ICA) within a 6-month period between 2011 and 2013, were included. Total perfusion deficit (TPD) was quantified automatically, and CAC scores from CTAC scans were assessed using artificial intelligence (AI)-derived segmentation and manual scoring. Obstructive CAD was defined as ≥50% stenosis in Left Main (LM) artery, or 70% or more stenosis in any of the other major epicardial vessels. Prediction performance for CAD was assessed by comparing the area under receiver operating characteristic curve (AUC) for stress TPD alone and in combination with CAC score.

Among 498 patients (72% males, median age 63 years) 30.1% had CAD. Incorporating CAC score resulted in a greater AUC: manual scoring (AUC=0.87, 95% Confidence Interval [CI] 0.34-0.90; p=0.015) and AI-based scoring (AUC=0.88, 95%CI 0.85-0.90; p=0.002) compared to stress TPD alone (AUC 0.84, 95% CI 0.80-0.92).

**Conclusions:** Combining automatically derived TPD and CAC score enhances ^18^F-flurpiridaz PET MPI accuracy in detecting significant CAD, offering a method that can be routinely used with PET/CT scanners without additional scanning or technologist time.

**CONDENSED ABSTRACT:** *Background:* We assessed the added value of CAC score from hybrid PET/CT CTAC scans combined with stress TPD for detecting significant CAD using novel ^18^F-flurpiridaz tracer

*Methods and results:* Patients from the ^18^F-flurpiridaz phase III clinical trial (n=498, 72% male, median age 63) who underwent PET/CT MPI and ICA within 6-months were included. TPD was quantified automatically, and CAC scores were assessed by AI and manual methods. Adding CAC score to TPD improved AUC for manual (0.87) and AI-based (0.88) scoring versus TPD alone (0.84).

*Conclusions:* Combining TPD and CAC score enhances ^18^F-flurpiridaz PET MPI accuracy for CAD detection

Graphical Abstract.
Overview of the study design.

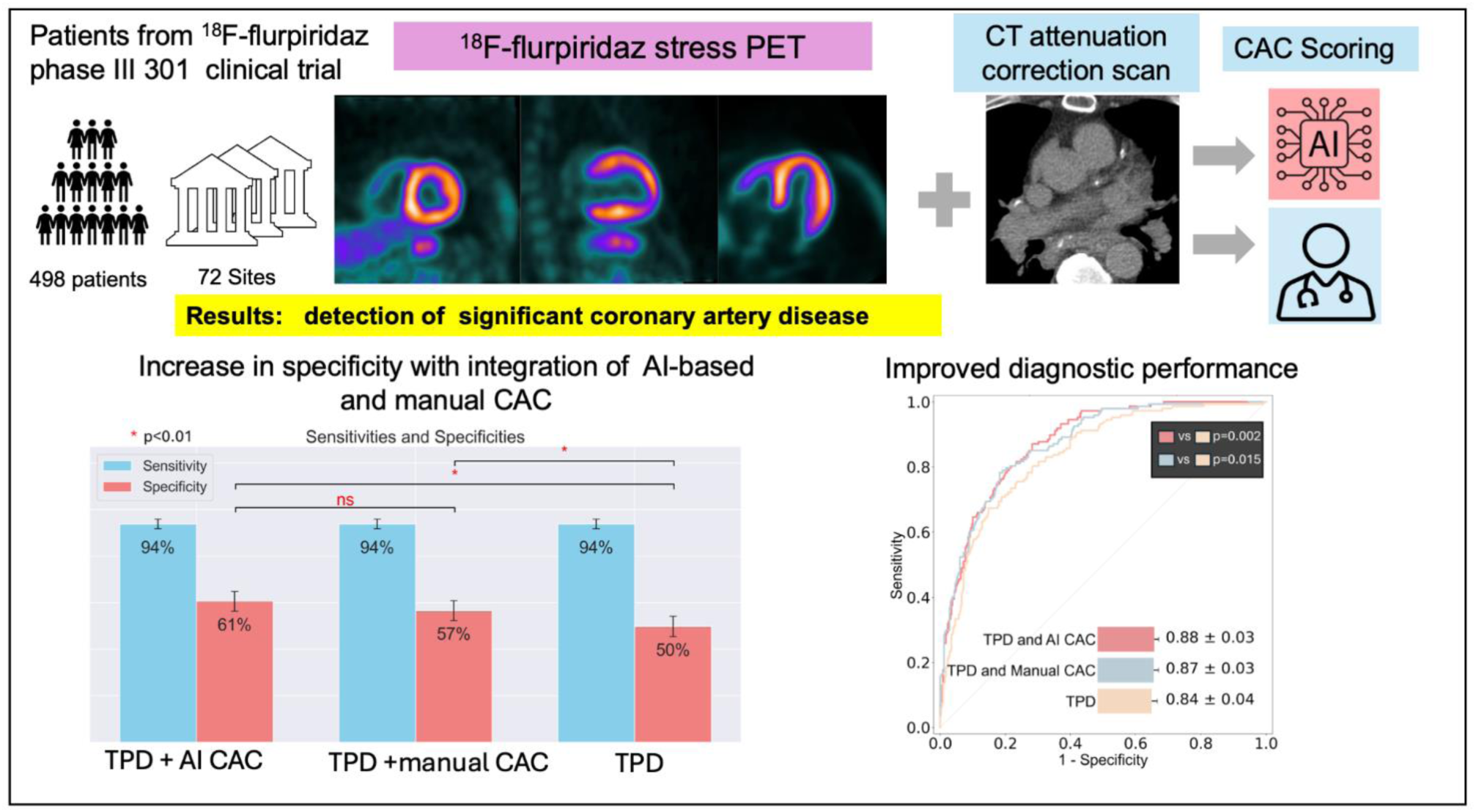

## INTRODUCTION

Positron Emission Tomography (PET) is the favored perfusion imaging technique according to the American Society of Nuclear Cardiology and Society of Nuclear Medicine and Molecular Imaging (2). However, its growth has been restricted due to logistic constraints and limitations associated with the currently available PET tracers, rubidium-82 (82Rb), nitrogen-13-ammonia (13N-ammonia), and oxygen-15-water (15O-water) (3,4). ^18^F-flurpiridaz is a novel PET-MPI radiotracer which has been demonstrated to be safe and effective in detecting CAD, offering superior image resolution compared to 82Rb, the most commonly used tracer, without the limitations associated with commonly used tracers. The ^18^F-flurpiridaz tracer was recently granted FDA regulatory approval and will be incorporated into routine clinical practice in the near future (5), potentially leading to significant growth in the use of PET scanning.

A coronary artery calcium (CAC) score is a powerful risk classifier for the prevalence of coronary artery disease (6). When combined with PET MPI, studies have shown that CAC score adds incremental risk stratification and enhances diagnostic accuracy(7–9). The introduction of hybrid PET/CT imaging techniques, along with the routine use of CTAC scans for attenuation correction, has enabled the assessment of CAC and MPI in a single scanning acquisition. New AI-based models have facilitated a fully automated quantification of CAC scores from CTAC imaging (10,11). While there is established data supporting the combination of CAC score and PET results using currently available tracers, whether incorporating the CAC score to PET-MPI using the novel radiotracer, ^18^F-flurpiridaz, will provide additional diagnostic value has not been studied.

We aimed to assess the diagnostic performance of automatically quantified stress TPD compared to stress TPD combined with CAC score from the CTAC scans, quantified either manually or through a deep learning-based technique as seen in the **Graphical Abstract.**

## METHODS

### Study population

Data from the first of the ^18^F-flurpiridaz phase III clinical trials (Lantheus Medical Imaging, NCT 01347710)(flurpiridaz-301), which enrolled 795 patients with known or suspected CAD who were referred for invasive coronary angiography (ICA) from 2011 to 2013 at 72 sites in the United States, Canada, and Finland, were retrospectively analysed(5). Patients with non-ischemic cardiomyopathy, a history of coronary artery bypass grafting or percutaneous coronary intervention within the previous 6 months, those with unstable cardiovascular conditions, and post heart transplantation patients were excluded (5).

The only exclusion criterion for this analysis was the unavailability of CTAC scans suitable for CAC scoring. Out of the 753 patient cases with evaluable stress PET and ICA data, 498 patients with available CTAC scans for CAC scoring were ultimately included in the analysis.

The study obtained IRB approval and complied with the Declaration of Helsinki. Written informed consent was obtained from all participants prior to their enrollment in the trial.

### PET/CT Protocol

In the Flurpiridaz-301 Trial, all patients underwent a rest/stress PET imaging acquisition sequence (5). Regadenoson, dipyridamole, or adenosine were used for pharmacological stress induction, and ^18^F-flurpiridaz was administered at doses of 2.5–3 mCi for rest, 6–6.5 mCi, with a minimum rest-stress dose interval of 30 minutes. For patients undergoing exercise stress, a dose of 9–9.5 mCi ^18^F-flurpiridaz was given with a minimum rest-exercise dose interval of 60 minutes (5,12). All PET-MPI studies performed with PET/CT include CTAC scans that are used for the AI-CAC assessment.

### Invasive coronary angiography procedure and coronary stenosis criteria

All participants underwent ICA within six months following the PET/CT MPI study. Each site conducted ICA according to local protocols before submitting the data to a core laboratory (PERFUSE Core Laboratories and Data Coordinating Center, Boston, Massachusetts) for quantitative analysis (5). Experienced interventional cardiologists visually analyzed and reported the results of the coronary angiograms. Consistent with widely accepted thresholds, significant CAD was defined was defined as ≥50% stenosis in the LM trunk or ≥70% stenosis in any other major epicardial coronary artery, including the LAD, the circumflex branch of the LCX, or the RCA, as determined by the ICA.

### Automated TPD quantification

Stress TPD, which reflects the extent and severity of myocardial perfusion defects, was quantified using QPET software (Cedars-Sinai, Los Angeles, California, United States) following the methods previously described (13,14). Per-vessel grouped stress TPD was computed separately for each vascular territory—LAD, LCX, and RCA regions based on the 17-segment model.

### Coronary artery calcium scoring methods

Standard per-vessel Agatston CAC scores and global (per-patient) CAC scores were assessed from the PET-CTAC maps and calculated separately for each participant using both manual scoring and a previously validated AI-based model (15). The agreement between deep learning (DL) and manual CAC quantification was assessed using linear weighted Cohen’s kappa coefficient and concordance values.

CAC scores for the LAD artery were calculated as the sum of the CAC scores of the LM artery and the LAD. CAC scores for the LCX artery were calculated as the sum of the CAC scores of the LM and the LCX.

The same CAC categories of 0, 1–99, 100–399, and 400 or greater were used for per-patient and per-vessel measures of the severity of CAC. These scores were used independently in conjunction with PET results for the prediction of CAD.

### Combined perfusion calcium models

Multivariable logistic regression was used to develop a quantitative score predicting obstructive CAD. To address the highly skewed distribution of CAC, we applied a log transformation as is conventional in studies assessing CAC in multivariable models (19).

The first combined model used stress TPD scores and per vessel DL-CAC scores. The second model used stress TPD scores and per vessel manual CAC scores. The prediction of obstructive CAD by the models were compared with the prediction by total stress TPD scores alone.

A 10-fold cross-validation (CV) technique (20) was utilized to ensure that none of the data used to train the logistic regression model was used in the subsequent testing of the diagnostic performance of the same model. This incurred the creation of 10 separate models each tested on distinct, randomly chosen “folds” containing 10% of the data, with the remaining data used for training. Results for prediction at each test set were aggregated and averaged, thereby effectively utilizing all patients in testing. The main benefit of utilizing this technique is the mitigation of overfitting. Creating 10 separate models each with varied training data makes for a more generalizable prediction than just 1 model.

### Diagnostic performance of PET versus hybrid approach

To assess differences in diagnostic performance for CAD detection between stress TPD alone versus stress TPD and CAC score, the area under the receiver operator characteristic curve (AUC) was used. Differences in diagnostic performance were evaluated in three feature groups: (1) univariate stress TPD alone, (2) grouped stress TPD and manual CAC score, and (3) grouped stress TPD and AI-based CAC score.

### Outcomes

The primary end point was the prediction of obstructive CAD based on ^18^F-flurpiridaz stress TPD results alone, compared to combined stress TPD and CAC score, as assessed by ICA results within six months.

### Statistical Analysis

Data is presented as mean ± standard deviation (SD) for normally distributed continuous variables, or median with interquartile range [IQR] for continuous variables with a skewed distribution. Categorical variables are reported as value counts. (20) Categorical variables were compared by using Pearson’s *χ*^2^ test, and continuous variables were compared by the student’s *t* test or Mann–Whitney U test, as appropriate. Concordance was assessed using Cohens Kappa scores. Significance in difference between ROC curves were calculated using a paired Delong test (21). The sensitivity and specificity of stress TPD, stress TPD with manual CAC score, and stress TPD with DL-CAC score were analyzed using McNemar’s test to compare differences in sensitivity and specificity between the methods. Sensitivity and specificity were initially calculated for univariate stress TPD using a standard 5% threshold. The resulting sensitivity was then applied to calculate specificity for the other two models. Additionally, reclassification analysis was conducted.

For each of the three major epicardial coronary vessels—LAD, LCX and RCA—we calculated CAC scores using both the automated DL model and manual annotation. The log-transformed CAC values, together with the global stress TPD, were used as input variables in a standard multivariable logistic regression model (Supplementary Figure 1) to predict the probability of obstructive coronary artery disease.

We developed two parallel versions of the multivariable logistic regression model: one using CAC values derived from the DL) algorithm and the other using manually annotated CAC scores. Both models were trained and evaluated using 10-fold cross-validation, ensuring that each patient was included in an independent test fold exactly once.

All statistical tests were two tailed and a p value<0.05 was considered significant. All statistical analyses were performed with Python 3.11.5 (Python Software Foundation, Wilmington, DE, USA).

## RESULTS

### Study Population

A total of patients 498 were included in the final analysis. **Table 1** summarizes patient characteristics for the entire cohort and according to the presence of significant CAD defined as ≥50% stenosis in LM trunk or ≥70% stenosis in LAD, LCX, or RCA. The median age of all participants was 63 (IQR 57,69) with 72.4% males. A total of 147 (30.1%) patients had confirmed CAD by ICA. Compared to patients without significant stenosis, those with significant CAD were older (65 years vs. 62 years), had a higher percentage of males (87% vs. 64%), and exhibited a greater prevalence of hyperlipidaemia (91.8% vs. 84.3%), diabetes mellitus (43.5% vs. 31.1%), prior PCI (42.9% vs. 24.8%), and smoking (66.7% vs. 55.3%). The calculated stress TPD was significantly higher in patients with detected stenosis (18.6 compared to 5.0; p<0.001). Similarly, both manually assessed, and DL-based CAC scores were markedly increased in patients with significant stenosis (650.5 vs. 44.9 and 538.5 vs. 22.4, respectively; p<0.001 for both).

**Table 1.** Patient characteristics.

|  | **All patients**<br>**N=498** | **Non-significant**<br>**Stenosis N=351** | **Significant stenosis**<br>**N=147** | **P Values** |
| --- | --- | --- | --- | --- |
| Age (median, IQR) | 63.0 (57.0, 69.0) | 62.0 (56.0, 68.0) | 65.0 (59.0, 71.0) | <0.001 |
| Sex, Male | 354 (71.1%) | 226 (64.4%) | 128 (87.1%) | <0.001 |
| BMI (median, IQR) | 30.0 (26.0, 34.0) | 30.0 (27.0, 34.5) | 29.0 (26.0, 33.0) | 0.114 |
| Hypertension | 404 (81.1%) | 282 (80.3%) | 122 (83.0%) | 0.573 |
| Hyperlipidemia | 431 (86.5%) | 296 (84.3%) | 135 (91.8%) | 0.042 |
| Diabetes Mellitus | 173 (34.7%) | 109 (31.1%) | 64 (43.5%) | 0.01 |
| Stroke | 31 (6.2%) | 20 (5.7%) | 11 (7.5%) | 0.583 |
| Myocardial Infarction | 73 (14.7%) | 43 (12.3%) | 30 (20.4%) | 0.039 |
| Prior PCI | 150 (30.1%) | 87 (24.8%) | 63 (42.9%) | <0.001 |
| Smoking | 292 (58.6%) | 194 (55.3%) | 98 (66.7%) | 0.021 |
| Family History of<br>CAD | 279 (56.0%) | 193 (55.0%) | 86 (58.1%) | 0.459 |
| **PET/CT results** |  |  |  |  |
| Exercise stress | 130 (26.1%) | 92 (26.2%) | 38 (25.9%) | 0.06 |
| Pharmacologic stress | 368 (73.9%) | 259 (76.6%) | 109 (74.1%) | 1.0 |
| TPD | 8.0 (3.46, 15.1) | 5.0(2.7,10.2) | 18.6(10.5, 25.1) | <0.001 |
| AI-based CAC score | 71.0 (0.0, 714.1) | 22.4 (0.0, 304.6) | 538.5 (79.4, 1525.3) | <0.001 |
| Manual CAC score | 42.5 (0.0, 753.4) | 44.9 (0.0, 389.7) | 650.5 (126.1, 1592.0) | <0.001 |

Values are expressed as the median (interquartile range) or number (%)
Body mass index (BMI), coronary artery disease (CAD), Total Perfusion Deficit (TPD),
Coronary Arteries Calcium (CAC), Artificial intelligence (AI)

**Table 2** displays the per vessel and per patient results of the ICA. Significant stenosis was observed in 147 (29.5%) of patients and was most prevalent in the RCA, followed by the LAD, LCX, and LM arteries. Approximately one-fifth of patients had significant stenosis in one vessel, while stenosis in two or three vessels was less frequently observed.

**Table 2.** Invasive coronary angiography results.

| **Vessel** | **Number of Patients** |
| --- | --- |
| No Stenosis | 351 (70.48%) |
| LAD Stenosis | 61 (12.25%) |
| LCX Stenosis | 55 (11.04%) |
| RCA Stenosis | 67 (13.45%) |
| LM Stenosis | 6 (1.20%) |
| 1 Vessel Stenosis | 111 (22.29%) |
| 2 Vessel Stenosis | 30 (6.02%) |
| 3 Vessel Stenosis | 6 (1.20%) |

**Supplementary Figures 2A–D**, illustrate the correlation between DL-derived and manually assessed CAC scores at both the total coronary tree and vessel-specific levels (LAD, RCA, and LCX). Each scatter plot includes both manual and DL-generated values, plotted on a log-transformed scale [log₁₀(CAC + 1)], along with a line of identity to visually assess agreement. Correlation was assessed using the Pearson correlation coefficient.

Figure 1A and 1B depict the prevalence of a significant CAD on a per-patient basis across the range of stress TPD and CAC scores quantified manually or by AI method (1A). Regarding stress TPD and manually calculated CAC scores, patients with TPD> 5% had the highest prevalence of significant stenosis on invasive angiography, with a stepwise increase in significant coronary stenosis across the categories of CAC scores. The presence of CAC >400 was associated with increased rate of obstructive CAD in patients in all TPD categories. Similar results were demonstrated for stress TPD and DL-based CAC scores (Figure 1B).

**Figure 1.**
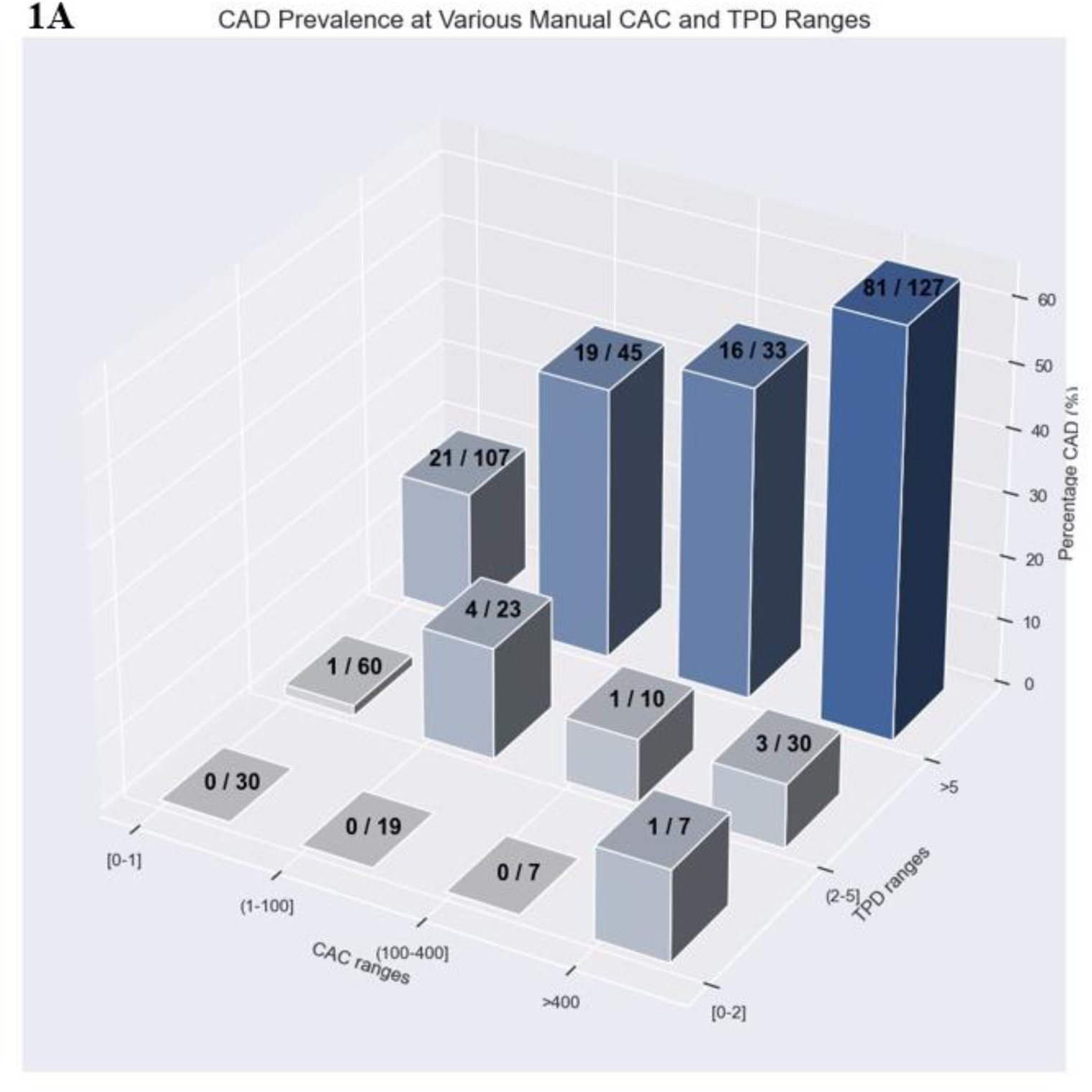
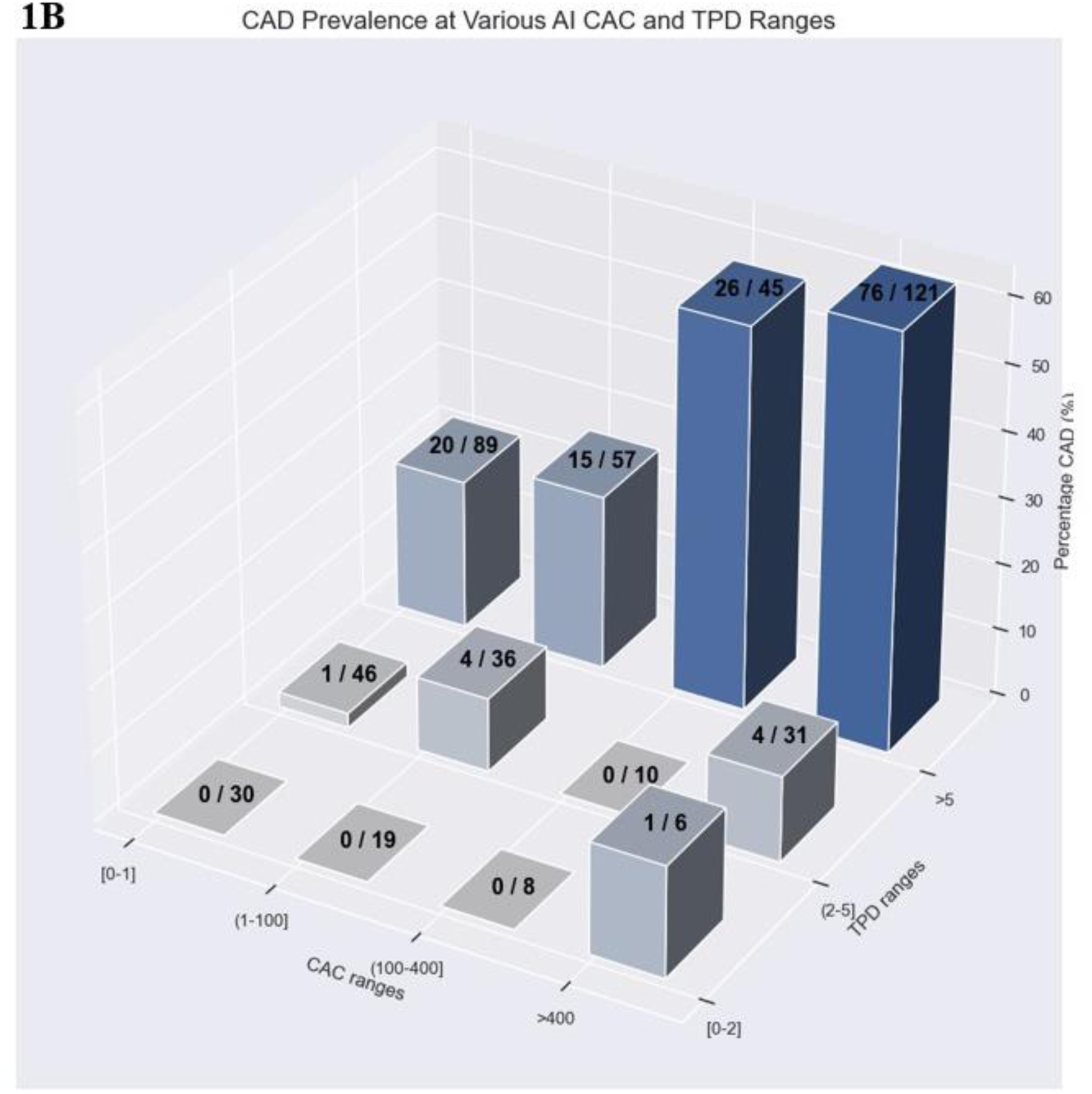
Prevalence of significant coronary artery disease across the range of stress total perfusion deficit (TPD) results and coronary artery calcium (CAC) scores quantified manually (1A) and by AI-based model(1B).

### Diagnostic performance Stress TPD alone and with the integration of CAC score

**Figure 2A and 2B** illustrate the diagnostic performance for significant CAD by incorporating CAC score into stress TPD results, compared to using stress TPD findings alone (2A) and the sensitivity and specificity for CAD detection when combining CAC score and stress TPD results compared to stress TPD alone (2B). Regarding model prediction performance (Figure 2A) the AUC for stress TPD combined with CAC score, calculated either manually (AUC 0.87, 95% CI 0.84-0.90; p=0.015) or by using AI technique (AUC 0.88, 95% CI 0.85-0.91; p=0.002) demonstrated greater performance for coronary stenosis prediction than that of TPD alone (AUC 0.84, 95% CI 0.80-0.92). AUC results for manually assessed and for DL-driven CAC score were comparable. Sensitivity and specificity for CAD detection using stress TPD results alone or combined with CAC score are shown in **Figure 2B wand Supplementary Figure 3**. Applying a threshold stress TPD of 5%, integrating the CAC score—whether derived from AI or manually calculated—with stress TPD results led to improved specificity (Figure 2B) but not sensitivity (**Supplementary Figure 3)** in detecting significant CAD over TPD alone. Additionally, in the reclassification analysis with specificity matched to the specificity of standard quantitative analysis, the number of detected true positives increased from 137 with TPD alone to 144 when combining Manual CAC+ TPD and DL-CAC + TPD.

**Figure 2.**
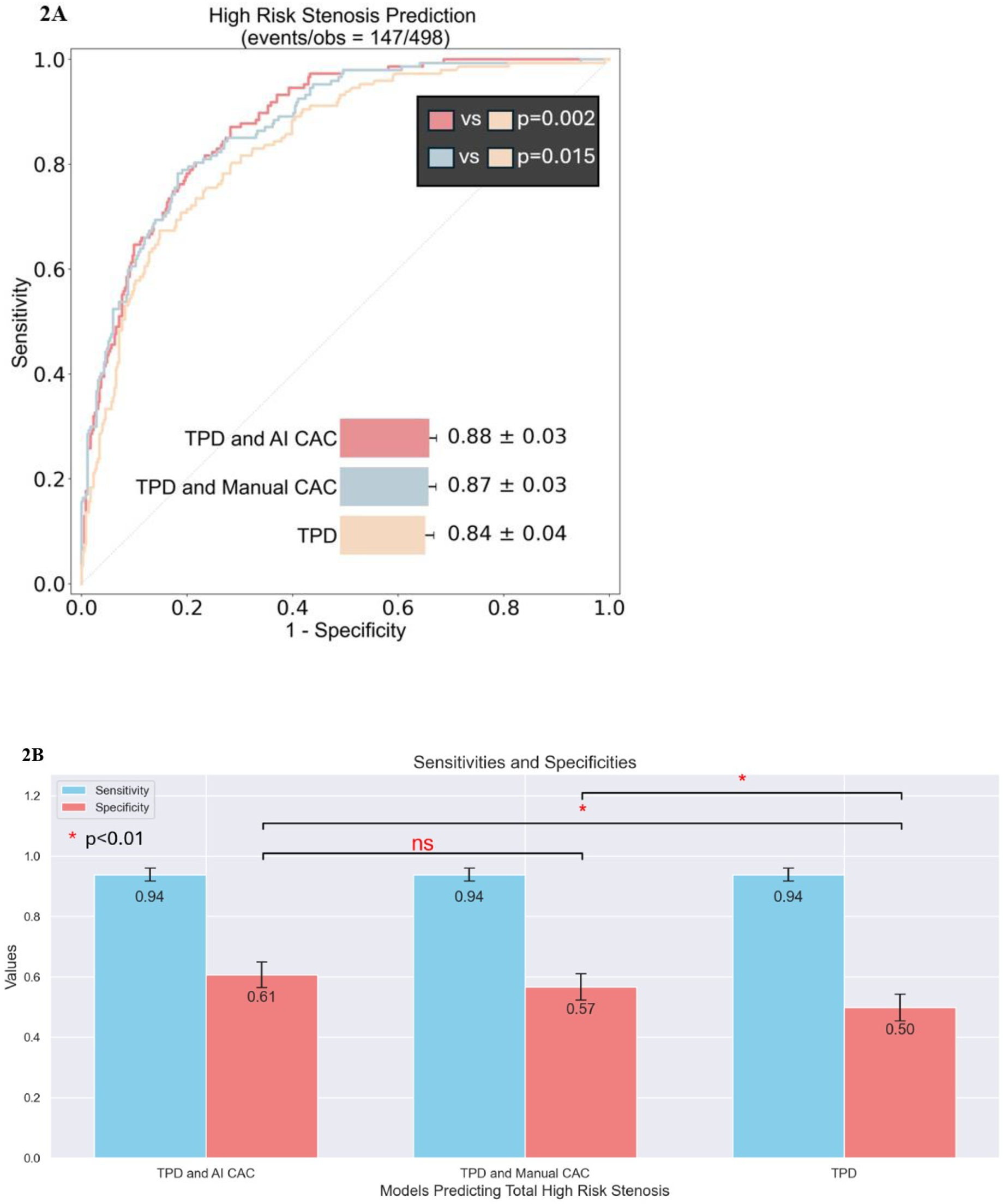
AUC (2A) with comparison to stress total perfusion deficit (TPD), Sensitivity (blue) and Specificity(red) (2B) for the detection of significant coronary artery disease (≥50% LM or ≥70% in other major epicardial arteries) of stress TPD combined with manual and AI-quantified CAC score compared with stress TPD alone. Sensitivity was derived from thresholding univariate stress TPD alone at >5% and matched across the 2 multivariate models. Specificities were calculated based on this matched sensitivity. There was significant increase in the specificity (p<0.001) for the AI-CAC addition as well in Manual CAC model (p=0.01) over TPD alone. AUC– area under the receiver operating characteristic curve Ns- non-significant

The diagnostic performance for significant CAD by incorporating CAC score into stress TPD results, compared to using stress TPD alone, was further analysed on a per-vessel basis, assessing the additive value of per-vessel CAC score for each coronary artery (Figure 3A-C). The results show that the CAC score, when quantified using an AI model, but not through manual assessment, provided a superior prediction for obstructive for the LCX artery compared to stress TPD alone (AUC 0.83 vs. 0.77; p=0.039). There was no incremental improvement with CAC in predicting significant CAD, for LAD or RCA arteries.

**Figure 3.**
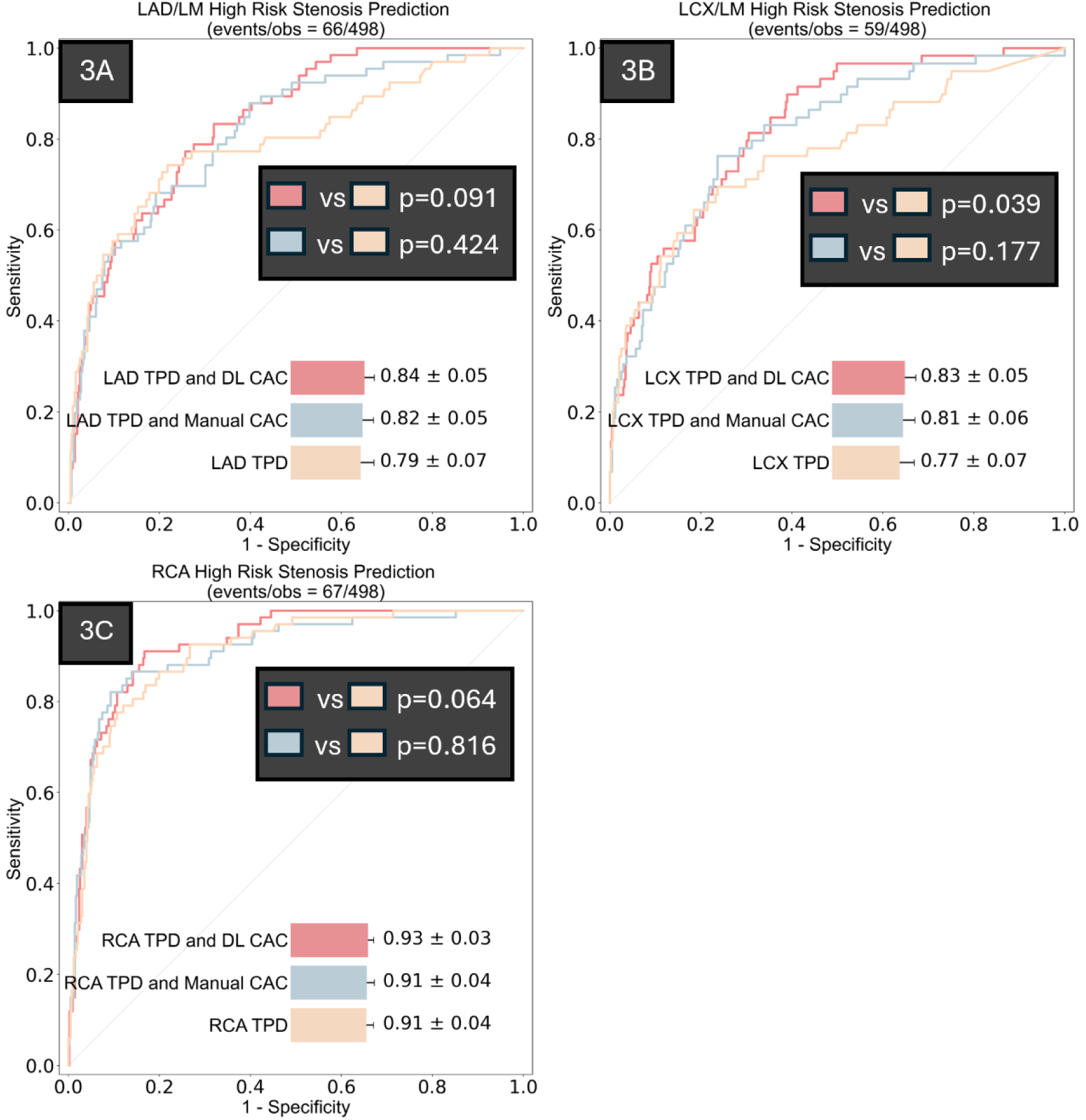
A-C. Per vessel assessment, A) LAD/LM; B) LCX/LM C) RCA, of model performance using AUC in predicting significant coronary artery disease comparing stress TPD combined with manual and DL CAC scores compared with stress TPD alone. AUC– area under the receiver operating characteristic curve, TPD-total perfusion deficit

## DISCUSSION

In this study we assessed the diagnostic performance for CAD detection of CAC score from CTAC maps in conjugation with automated quantitative stress TPD, for ^18^F-flurpiridaz PETMPI. Accordingly, we analyzed 498 patients from the ^18^F-flurpiridaz trial who underwent hybrid PET/CT MPI with the utilization of CTAC maps and had an ICA withing 6 months.

Incorporating CAC scores into the TPD findings improved the accuracy of obstructive CAD prediction compared to TPD alone. Findings were comparable for both DL-CAC and manual CAC and demonstrated that CAC score provides superior diagnostic accuracy for obstructive CAD prediction when combined with high-resolution PET tracers such as ^18^F-flurpiridaz.

Additionally, we assessed the value of the predictive model per specific vessel. The per-vessel analysis is clinically relevant, as disease in different coronary arteries may have varying prognostic implications and diagnostic challenges. Our results showed variation in model performance across vessels, likely reflecting both anatomical and technical factors—such as differences in vessel size, motion artifacts, and calcification patterns—that influence image quality and interpretability.

Incorporating vessel-specific CAC improved the ability to localize disease and added diagnostic value beyond stress TPD alone. This supports its potential utility in refining risk assessment and guiding more targeted clinical decision-making.

With the anticipated introduction of 18F-flurpiridaz into clinical practice, our findings emphasize the significance of incorporating CAC burden alongside results from ^18^F-flurpiridaz PET-MPI to enhance prediction of significant CAD.

The additive value of CAC score to PET-MPI in diagnosing CAD was previously reported. In the study of Thompson et al. (22) it was demonstrated that 17% of patients with non-ischemic MPI were reclassified as having significant coronary atherosclerosis based on a CAC score exceeding 100. Similarly, Schepis et al. (23) showed that combining CAC score at a cutoff of 709 to MPI further identifies patients with significant CAD who were missed by MPI. These findings were further supported by a Ghadri et al. (24) study which showed that a CAC score of 1,000 or greater provides significant diagnostic value for detecting significant coronary artery disease, in patients with normal MPI results. Brodov et al. (9) evaluated the predictive accuracy of obstructive CAD for any combination of ITPD and CAC values and found that combining automatically derived per-vessel ITPD results with the logCAC score significantly improved the accuracy of 82Rb PET MPI in detecting obstructive CAD. Schenker et al. (8) and Chang et al. (25) demonstrated a stepwise increase in risk of adverse cardiac events with increasing CAC in patients with and without ischemia. Consistent with the aforementioned studies, our study confirms an improvement in CAD detection when combining CAC score with ^18^F-flurpiridaz PET-MPI results.

The recently FDA-approved ^18^F-flurpiridaz PET-MPI radiotracer offers improved image quality and enhances diagnostic accuracy for detecting CAD (5). To our knowledge, the role of CAC score in the context of tracers with high sensitivity for detecting milder perfusion defects, such as 13N ammonia and ^18^F-flurpiridaz, which allow for a more accurate evaluation of the true extent of perfusion abnormalities, has not been previously assessed. Our study demonstrates that in patients with known or suspected CAD undergoing hybrid PET/CT imaging with the ^18^F-flurpiridaz, incorporating CAC scores from CTAC maps alongside stress TPD findings significantly enhances the prediction of significant CAD. This approach improved both the sensitivity and specificity of CAD detection, with an increase in diagnostic performance compared to stress TPD alone.

For CAC quantification, we analyzed the CT images used for attenuation correction in hybrid PET/CT scans. Although these images are not routinely employed for this purpose, PET CTAC maps can provide a reliable quantitative assessment of CAC (26). The advantage of using CTAC images for CAC assessment is that it eliminates the need for separate imaging acquisitions, thus, reduces radiation exposure, minimizes exam time. Using -based CAC quantification from CT imaging in hybrid PET/CT MPI yields similar risk stratification to standard expert CAC scores while eliminating the time required for manual scoring (11,26).

While the level of agreement between AI-based and manual CAC quantification, assessed using Cohen’s kappa coefficient and concordance, was influenced by factors such as low CTAC image quality and the presence of intracoronary stents, we demonstrated strong concordance between the AI-based and manual approaches, with excellent correlations observed for total CAC score (r = 0.85), as well as for individual vessels: LAD (r = 0.89), RCA (r = 0.84), and LCX (r = 0.77). supporting the robustness and clinical utility of the AI model in reliably quantifying CAC across the major coronary arteries.

CAC was modelled as a continuous, log-transformed variable, allowing the model to capture the full spectrum of CAC burden without requiring a predefined Agatston score threshold.

Using this model, our results demonstrate a comparable improvement in CAD prediction by integrating PET-MPI results with the CTAC CAC score—whether quantified manually or analyzed using an AI model. Similarly, both manually quantified CAC and AI-derived CAC score demonstrated higher sensitivity and specificity for identifying significant CAD compared to stress PET-MPI findings alone.

Of note, the sensitivities for detecting CAD using TPD alone and in combination with CAC in our study were higher than those reported in the original paper. This is likely due to the greater sensitivity of the quantitative TPD measurements used in our analysis compared to the visual interpretations applied in the overall Flurpiridaz-301 trial. Additionally, whereas prior literature has largely emphasized improvements in sensitivity with hybrid CAC and perfusion imaging approaches, our analysis demonstrated the observed increase in AUC. Depending on the abnormality threshold chosen we can demonstrate increase in sensitivity or specificity. The improvement in specificity is related to the very low likelihood of obstructive CAD in patients with no or mild CAC scores. These findings highlight the value of CAC in enhancing diagnostic precision—particularly by more accurately identifying true negatives and reducing false-positive results.

### Conclusion

The accuracy of CAD detection using stress TPD findings from ^18^F-flurpiridaz PET MPI is significantly enhanced by the inclusion of quantified CAC burden. The findings of this study underscore the importance of employing integrated anatomical and functional imaging techniques in clinical practice to more accurately identify patients at risk for obstructive CAD, even when using more advanced PET MPI tracers like ^18^F-flurpiridaz.

### Clinical implications

Due to its nearly ideal characteristics as a myocardial perfusion tracer and its ability to enable PET-MPI on low-volume PET sites, ^18^F-flurpiridaz PET-MPI holds promise as one of the most widely used tracers for PET MPI.

Our results indicate that integrating manual or AI-derived CTAC CAC scores with quantitative TPD analysis in stress PET studies can better identify patients with significant CAD than relying solely on ^18^F-flurpiridaz PET-MPI findings. The automation of CAC and stress TPD results will facilitate easy integration into clinical routines. and enable rapid, highly sensitive assessments for the presence of significant CAD.

### Limitations

Our study has several limitations. First, our study population includes data from the first of the ^18^F-flurpiridaz phase III clinical trials (Lantheus Medical Imaging, NCT 01347710) (flurpiridaz-301) which enrolled patients with suspected or pre-existing coronary artery disease. The efficacy of automated CAC scoring in patients with pre-existing CAD and coronary stents remains uncertain. Second, our inclusion criteria—specifically the requirement for both PET stress TPD and available CTAC scans to compute CAC scores—may have introduced selection bias. Patients included in this cohort may have had higher-quality imaging, as only those with adequate CTAC scans were eligible, potentially resulting in better image quality compared to the overall study population. This selection bias could have led to an attenuation of sensitivity estimates, as patients with non-interpretable or missing data (who may have different disease profiles) were under-represented. Third, CT imaging quality and the presence of intracoronary stents, pacemaker clips may have impacted the level of agreement between AI-driven and manually assessed CAC. However, good agreement between the two methods was still observed, and both demonstrated comparable performance in predicting CAD.

### Sources of funding

This research was supported in part by grant R35HL161195 from the National Heart, Lung, and Blood Institute at the National Institutes of Health and R01EB034586 from the National Institute of Biomedical Imaging and Bioengineering of the National Institutes of Health. This work was also supported in part by the Dr. Miriam and Sheldon G. Adelson Medical Research Foundation and a grant from the American College of Cardiology in support of the Asher Kimchi Memorial Fellowship. The content is solely the responsibility of the authors and does not necessarily represent the official views of the National Institutes of Health.

## Data Availability

To the extent allowed by data sharing agreements and IRB protocols, the deidentified data and data analysis code from this manuscript will be shared upon written request.

## Abbreviations

PET: Positron Emission Tomography
MPI: Myocardial Perfusion Imaging
TPD: Total Perfusion Deficit
CTAC: Coronary Tomography Attenuation Correction
CAC: Coronary Artery Calcium
ICA: Invasive Coronary Angiography
AI: Artificial Intelligence
CAD: coronary artery disease

