## Supplemental Material for "Incremental diagnostic value of AI-derived coronary artery calcium in ^18^F-flurpiridaz PET Myocardial Perfusion Imaging"

**SUPPLEMENTAL MATERIALS**

**Supplementary Figure 1.** Multivariable logistic regression model

$$\log\left(\frac{p}{1-p}\right) = \beta_0 + \beta_1(TPD) + \beta_2(LAD\ LM\ CAC) + \beta_3(LCX\ LM\ CAC) + \beta_4(RCA\ CAC)$$

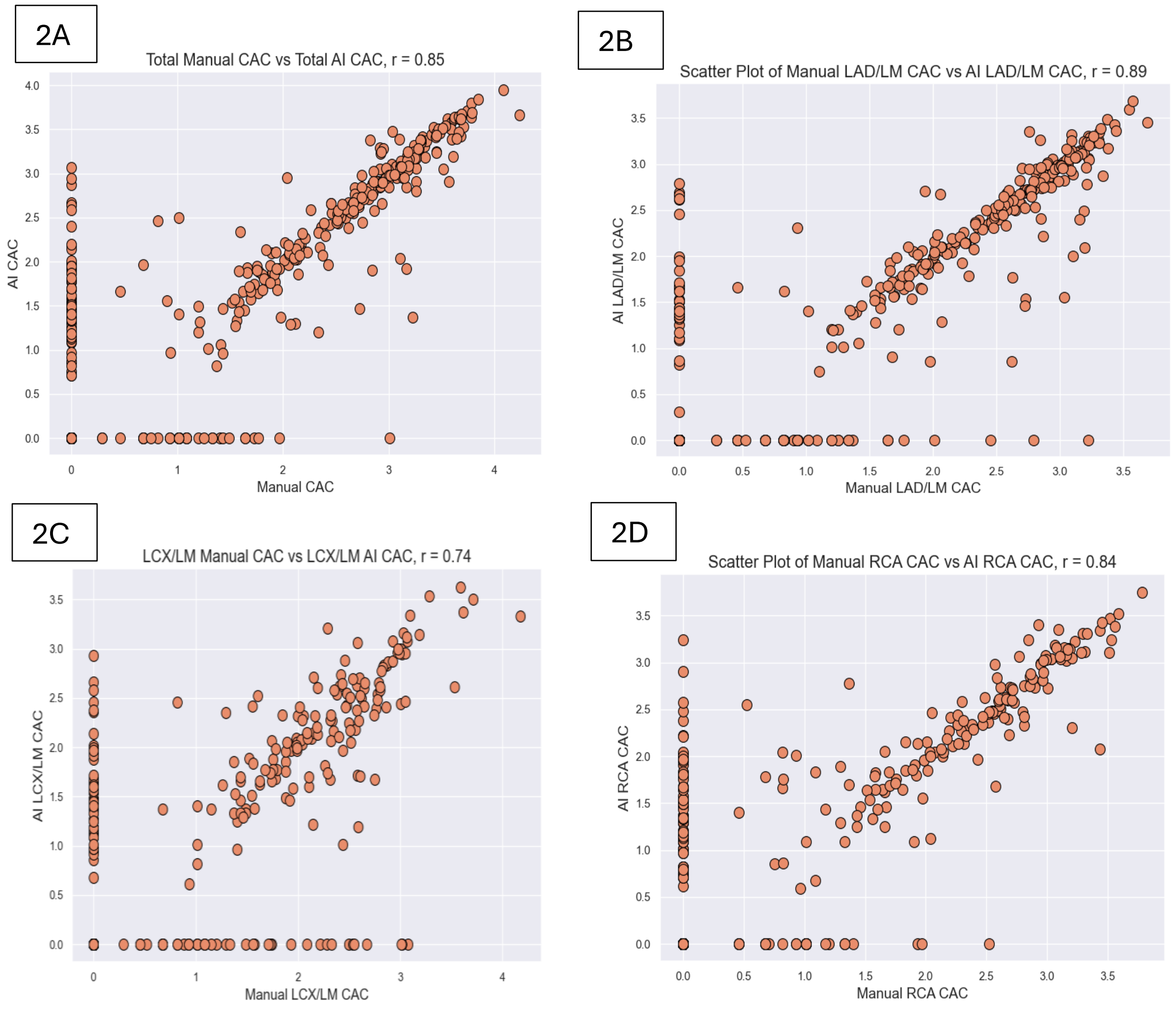


**Supplementary Figure 2A-D**, illustrate the correlation between AI-derived and manually assessed CAC scores at both the total coronary tree (2A) and vessel-specific levels- LAD/LM (2B), LCX/LM (2C) and RCA (2D), and. Each scatter plot includes both manual and AI-generated values, plotted on a log-transformed scale [$\log_{10}$(CAC + 1)], along with a line of identity to visually assess agreement. Correlation was assessed using the Pearson correlation coefficient.

Artificial Intelligence (AI), Coronary Artery Calcification (CAC), Left Main (LM), Left Anterior Descending (LAD), Left Circumflex (LCX), Right Coronary Artery (RCA)

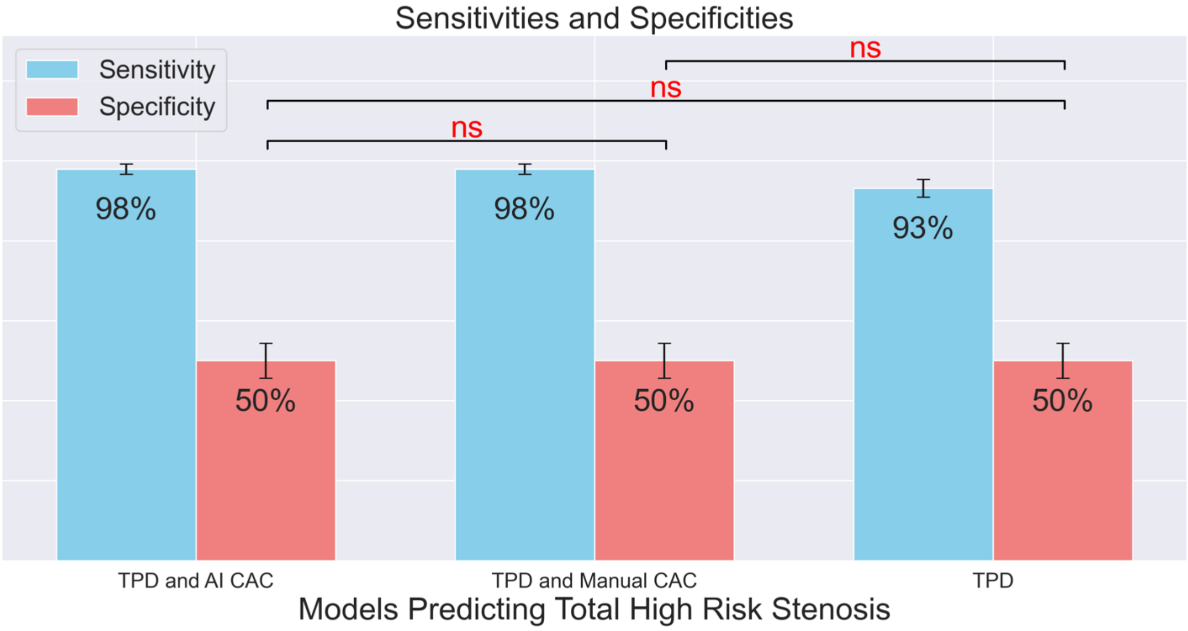


**Supplementary Figure 3.** Sensitivity (blue) and Specificity (red) for the detection of significant coronary artery disease (≥50% LM or ≥70% in other major epicardial arteries) of stress TPD combined with manual and AI-quantified CAC score compared with stress TPD alone. Specificity was derived from thresholding univariate stress TPD alone at >5% and matched across the 2 multivariate models. Sensitivities were calculated based on this matched Specificity. There was no significant increase in the sensitivity for the AI CAC addition as well in Manual CAC model over TPD alone.

Ns- non-significant

Artificial Intelligence (AI), Coronary Artery Calcification (CAC), Total Perfusion Deficit (TPD)
